## Supplementary tables and figures for "Definitions matter: heterogeneity of COVID-19 disease severity criteria and incomplete reporting compromise meta-analysis"

Supplementary Table 1. Severity definitions

|  | **WHO - Clinical management of COVID-19^a^** | **WHO - Therapeutics and COVID-19: living guideline^b^** | **NIH - COVID-19 Treatment Guidelines^c^** | **COVID-19 Living Network Meta-Analysis group^d^** | **COVID NMA initiative^e^** |
| --- | --- | --- | --- | --- | --- |
| Asymptomatic/pre-symptomatic |  | Non-severe: absence of any signs of severe of critical disease | Individuals who test positive for SARS-CoV-2 using a virologic test (i.e., a nucleic acid amplification test or an antigen test) but who have no symptoms that are consistent with COVID-19 |  |  |
| Mild | Symptomatic patients meeting the case definition for COVID-19 without evidence of viral pneumonia or hypoxia |  | Individuals who have any of the various signs and symptoms of COVID-19 (e.g., fever, cough, sore throat, malaise, headache, muscle pain, nausea, vomiting, diarrhoea, loss of taste and smell) but who do not have shortness of breath, dyspnoea, or abnormal chest imaging | Clinical symptoms are mild with no sign of pneumonia on imaging | Mild disease ambulatory:  "outpatients" whose clinical symptoms are mild with no sign of pneumonia on imaging  OR  Mild disease:  clinical symptoms requiring hospitalization but no need for supplemental oxygen |
| Moderate | Adolescent or adult with clinical signs of pneumonia (fever, cough, dyspnoea, fast breathing) but no signs of severe pneumonia, including SpO2≥ 90% on room air |  | Individuals who show evidence of lower respiratory disease during clinical assessment or imaging and who have saturation of oxygen (SpO_2_) ≥94% on room air at sea level | Patients have fever and respiratory symptoms with radiological findings of pneumonia and requiring oxygen (3L/min>Oxygen <5L/min) | Fever and respiratory symptoms with radiological findings of pneumonia and requiring standard oxygen therapy O2 (3–5 L/min) |
| Severe | Adolescent or adult with clinical signs of pneumonia (fever, cough, dyspnoea, fast breathing) plus one of the following: respiratory rate > 30 breaths/min; severe respiratory distress; or SpO2 < 90% on room air | Oxygen saturation <90% on room air  OR  Respiratory rate >30 breaths per minute  OR  Signs of severe respiratory distress (accessory muscle use, inability to complete full sentences) | Individuals who have SpO_2_ <94% on room air at sea level, a ratio of arterial partial pressure of oxygen to fraction of inspired oxygen (PaO_2_/FiO_2_) <300 mm Hg, respiratory frequency >30 breaths/min, or lung infiltrates >50% | Fever or suspected respiratory infection, AND  Respiratory rate > 30 breaths/min  OR  Severe respiratory distress  OR  Arterial oxygen saturation (SpO2) ≤ 93% on room air. | Respiratory distress (≧ 30 breaths/min)  OR  Oxygen saturation ≤ 93% at rest in ambient air or oxygen saturation ≤97% with O2 > 5 L/min  OR  PaO2/FiO2 ≦ 300 mmHg (l mmHg=0.133 kPa). PaO2/FiO2 in high-altitude areas (> 1000 m above sea level) is corrected by the following formula: PaO2/FiO2 x [atmospheric pressure (mmHg)/760]  OR  Patients hospitalized on Non-invasive ventilation (NIV)/High flow Nasal oxygen (HFNO) |
| Critical | Acute respiratory distress syndrome (ARDS) defined by chest imaging (radiograph, CT scan, or lung ultrasound) with bilateral opacities, not fully explained by volume overload, lobar or lung collapse, or nodules.  AND  Oxygenation impairment in adults: Mild ARDS: 200 mmHg < PaO2/FiO2a≤ 300 mmHg (with PEEP or CPAP ≥ 5 cmH2O)  Moderate ARDS: 100 mmHg < PaO2/FiO2≤ 200 mmHg (with PEEP ≥ 5 cmH2O)  Severe ARDS: PaO2/FiO2≤ 100 mmHg (with PEEP ≥ 5 cmH2O)  OR  Sepsis defined by acute life-threatening organ dysfunction caused by a dysregulated host response to suspected or proven infection. Signs of organ dysfunction include: altered mental status, difficult or fast breathing, low oxygen saturation, reduced urine output, fast heart rate, weak pulse, cold extremities or low blood pressure, skin mottling, laboratory evidence of coagulopathy, thrombocytopenia, acidosis, high lactate, or hyperbilirubinemia  OR  Septic shock defined by persistent hypotension despite volume resuscitation, requiring vasopressors to maintain MAP ≥ 65mmHg and serum lactate level > 2 mmol/L | Defined by the criteria for acute respiratory distress syndrome (ARDS), sepsis, septic shock, or other conditions that would normally require the provision of life sustaining therapies such as mechanical ventilation (invasive or non-invasive) or vasopressor therapy | Individuals who have respiratory failure, septic shock, and/or multiple organ dysfunction | Presence of ARDS  AND/OR  an expected survival of <48 hours | Cases meeting any of the following criteria:  Respiratory failure requiring invasive mechanical ventilation  OR  Shock  OR  Other organ failure requiring Intensive Care Unit care |

^a^ <https://apps.who.int/iris/handle/10665/332196>

^b^ <https://apps.who.int/iris/handle/10665/340374>

^c^ <https://www.covid19treatmentguidelines.nih.gov/overview/clinical-spectrum/>

^d^ <https://www.covid19lnma.com/drug-treatments-study-level/> Definitions available in “about this table” (15 April 2021)

^e^ <https://covid-nma.com/living_data/more_details.php> (10 April 2021)
