## Supplementary tables and figures for "Definitions matter: heterogeneity of COVID-19 disease severity criteria and incomplete reporting compromise meta-analysis"

Supplementary Table 2. Trial publication details

| **Publication abbreviation** | **Citation** | **DOI** | **Trial registry ID as reported in publication** |
| --- | --- | --- | --- |
| Abbaspour Kasgari 2020 | Abbaspour Kasgari H, Moradi S, Shabani AM, et al. Evaluation of the efficacy of sofosbuvir plus daclatasvir in combination with ribavirin for hospitalized COVID-19 patients with moderate disease compared with standard care: a single-centre, randomized controlled trial. Journal of Antimicrobial Chemotherapy. 2020 Nov;75(11):3373-8. | 10.1093/jac/dkaa332 | IRCT20200328046886N1 |
| Abd-Elsalam 2020 | Abd-Elsalam S, Esmail ES, Khalaf M, et al. Hydroxychloroquine in the treatment of COVID-19: a multicenter randomized controlled study. The American Journal of Tropical Medicine and Hygiene. 2020 Oct 7;103(4):1635-9. | 10.4269/ajtmh.20-0873 | NCT04353336 |
| Altay 2020 | Altay O, Yang H, Aydin M, et al. Combined metabolic cofactor supplementation accelerates recovery in mild-to-moderate COVID-19. medRxiv. 2020.10.02.20202614 | 10.1101/2020.10.02.20202614 | NCT04573153 |
| Angus 2020 | Angus DC, Derde L, Al-Beidh F, et al. Effect of hydrocortisone on mortality and organ support in patients with severe COVID-19: the REMAP-CAP COVID-19 corticosteroid domain randomized clinical trial. Jama. 2020 Oct 6;324(13):1317-29. | 10.1001/jama.2020.17022 | NCT02735707 |
| Ansarin 2020 | Ansarin K, Tolouian R, Ardalan M, et al. Effect of bromhexine on clinical outcomes and mortality in COVID-19 patients: A randomized clinical trial. BioImpacts: BI. 2020;10(4):209. | 10.34172/bi.2020.27 | IRCT202003117046797N4 |
| Beigel 2020 | Beigel JH, Tomashek KM, Dodd LE, et al. Remdesivir for the treatment of Covid-19-preliminary report. The New England journal of medicine. 2020 May 22. | 10.1056/NEJMoa2007764 | NCT04280705 |
| Cao 2020a | Cao B, Wang Y, Wen D, et al. A trial of lopinavir-ritonavir in adults hospitalized with severe Covid-19. New England Journal of Medicine. 2020 Mar 18. | 10.1056/NEJMoa2001282 | ChiCTR2000029308 |
| Cao 2020b | Cao Y, Wei J, Zou L, et al. Ruxolitinib in treatment of severe coronavirus disease 2019 (COVID-19): A multicenter, single-blind, randomized controlled trial. Journal of Allergy and Clinical Immunology. 2020 Jul 1;146(1):137-46. | 10.1016/j.jaci.2020.05.019 | NA |
| Castillo 2020 | Castillo ME, Costa LM, Barrios JM, et al. Effect of calcifediol treatment and best available therapy versus best available therapy on intensive care unit admission and mortality among patients hospitalized for COVID-19: A pilot randomized clinical study. The Journal of steroid biochemistry and molecular biology. 2020 Oct 1;203:105751. | 10.1016/j.jsbmb.2020.105751 | NCT04366908 |
| Cavalcanti 2020 | Cavalcanti AB, Zampieri FG, Rosa RG, et al. Hydroxychloroquine with or without Azithromycin in Mild-to-Moderate Covid-19. New England Journal of Medicine. 2020 Nov 19;383(21):2041-52. | 10.1056/NEJMoa2019014 | NCT04322123 |
| Chen 2020a | Chen Z, Hu J, Zhang Z et al. Efficacy of hydroxychloroquine in patients with COVID-19: results of a randomized clinical trial. medrxiv. 2020.03.22.20040758 | 10.1101/2020.03.22.20040758 | ChiCTR2000029559 |
| Chen 2020b | Chen C, Huang J, Cheng Z, et al. P. Favipiravir versus arbidol for COVID-19: a randomized clinical trial. medRxiv. 2020.03.17.20037432 | 10.1101/2020.03.17.20037432 | ChiCTR2000030254 |
| Chen 2020d | Chen L, Zhang ZY, Fu JG, et al. Efficacy and safety of chloroquine or hydroxychloroquine in moderate type of COVID-19: a prospective open-label randomized controlled study. medRxiv. 2020.06.19.20136093 | 10.1101/2020.06.19.20136093 | ChiCTR2000030054 |
| Chen 2020e | Chen CP, Lin YC, Chen TC et al. A Multicenter, randomized, open-label, controlled trial to evaluate the efficacy and tolerability of hydroxychloroquine and a retrospective study in adult patients with mild to moderate Coronavirus disease 2019 (COVID-19). PloS one. 2020 Dec 2;15(12):e0242763. | 10.1371/journal.pone.0242763 | NCT04384380 |
| Cheng 2020 | Cheng LL, Guan WJ, Duan CY, et al. Effect of Recombinant Human Granulocyte Colony-Stimulating Factor for Patients With Coronavirus Disease 2019 (COVID-19) and Lymphopenia: A Randomized Clinical Trial. JAMA Internal Medicine. 2021 Jan 1;181(1):71-8. | 10.1001/jamainternmed.2020.5503 | ChiCTR2000030007 |
| Corral-Gudino 2020 | Corral-Gudino L, Bahamonde A, Arnaiz-Revillas F et al. Methylprednisolone in adults hospitalized with COVID-19 pneumonia. Wien Klin Wochenschr. 2021 133, 303–311 | 10.1007/s00508-020-01805-8 | EUCTR2020-001934-37 |
| Cruz 2020 | Cruz LR, Baladron I, Rittoles A, et al. Treatment with an Anti-CK2 Synthetic Peptide Improves Clinical Response in Covid-19 Patients with Pneumonia. A Randomized and Controlled Clinical Trial. ACS Pharmacol Transl Sci. 2020 Dec 11;4(1):206-212 | 10.1021/acsptsci.0c00175 | RPCEC00000317 |
| Dabbous 2020 | Dabbous HM, El-Sayed MH, El Assal G, et al. A Randomized Controlled Study Of Favipiravir Vs Hydroxychloroquine In COVID-19 Management: What Have We Learned So Far? Research square 2020; doi: 10.21203/rs.3.rs-83677/v1 | 10.21203/rs.3.rs-83677/v1 | NCT04349241 |
| Davoodi 2020 | Davoodi L, Abedi SM, Salehifar E, et al. Febuxostat therapy in outpatients with suspected COVID-19: A clinical trial. International journal of clinical practice. 2020 Nov;74(11):e13600. | 10.1111/ijcp.13600 | IRCT2019072704434N1 |
| Davoudi-Monfared 2020 | Davoudi-Monfared E, Rahmani H, Khalili H, et al. A Randomized Clinical Trial of the Efficacy and Safety of Interferon β-1a in Treatment of Severe COVID-19. Antimicrob Agents Chemother. 2020 Aug 20;64(9):e01061-20. | 10.1128/AAC.01061-20 | IRCT20100228003449N28 |
| Deftereos 2020 | Deftereos SG, Giannopoulos G, Vrachatis DA, et al. Effect of colchicine vs standard care on cardiac and inflammatory biomarkers and clinical outcomes in patients hospitalized with coronavirus disease 2019: the GRECCO-19 randomized clinical trial. JAMA network open. 2020 Jun 1;3(6):e2013136-. | 10.1001/jamanetworkopen.2020.13136 | NCT04326790 |
| Delgado-Enciso 2020 | Delgado-Enciso I, Paz-Garcia J, Barajas-Saucedo CE, et al. Patient-Reported Health Outcomes After Treatment of COVID-19 with Nebulized and/or Intravenous Neutral Electrolyzed Saline Combined with Usual Medical Care Versus Usual Medical care alone: A Randomized, Open-Label, Controlled Trial. Research Square 2020; doi:10.21203/rs.3.rs-68403/v1 | 10.21203/rs.3.rs-68403/v1 | RPCEC00000309 |
| Dequin 2020 | Dequin PF, Heming N, Meziani F, et al. Effect of hydrocortisone on 21-day mortality or respiratory support among critically ill patients with COVID-19: a randomized clinical trial. Jama. 2020 Oct 6;324(13):1298-306. | 10.1001/jama.2020.16761 | NCT02517489 |
| Doi 2020 | Doi Y, Hibino M, Hase R, et al. A prospective, randomized, open-label trial of early versus late favipiravir therapy in hospitalized patients with COVID-19. Antimicrob agents chemother. 2020 Nov 17;64(12). | 10.1128/AAC.01897-20 | jRCTs041190120 |
| Duarte 2020 | Duarte M, Pelorosso FG, Nicolosi L, et al. Telmisartan for treatment of Covid-19 patients: an open randomized clinical trial. Preliminary report. medRxiv. 2020.08.04.20167205 | 10.1101/2020.08.04.20167205 | NCT04355936 |
| Edalatifard 2020 | Edalatifard M, Akhtari M, Salehi M, et al. Intravenous methylprednisolone pulse as a treatment for hospitalised severe COVID-19 patients: results from a randomised controlled clinical trial. European Respiratory Journal. 2020 Dec 1;56(6). | 10.1183/13993003.02808-2020 | IRCT20200404046947N1 |
| Esquivel-Moynelo 2020 | Idelsis EM, Jesus PE, Yaquelin DR, et al. Effect and safety of combination of interferon alpha-2b and gamma or interferon alpha-2b for negativization of SARS-CoV-2 viral RNA. Preliminary results of a randomized controlled clinical trial. medRxiv. 2020.07.29.20164251 | 10.1101/2020.07.29.20164251 | RPCEC00000307 |
| Farahani 2020 | Farahani RH, Mosaed R, Nezami-Asl A, et al. Evaluation of the efficacy of methylprednisolone pulse therapy in treatment of covid-19 adult patients with severe respiratory failure: randomized, clinical trial. Research Square. 2020. https://doi.org/10.21203/rs.3.rs-66909/v1 | 10.21203/rs.3.rs-66909/v1 | IRCT20200406046963N1 |
| Fu 2020 | Fu W, Liu Y, Liu L, et al. An open-label, randomized trial of the combination of IFN-Îº plus TFF2 with standard care in the treatment of patients with moderate COVID-19. EClinicalMedicine. 2020 Oct 1;27:100547. | 10.1016/j.eclinm.2020.100547 | ChiCTR2000030262 |
| Furtado 2020 | Furtado RH, Berwanger O, Fonseca HA, et al. Azithromycin in addition to standard of care versus standard of care alone in the treatment of patients admitted to the hospital with severe COVID-19 in Brazil (COALITION II): a randomised clinical trial. The Lancet. 2020 Oct 3;396(10256):959-67. | 10.1016/S0140-6736(20)31862-6 | NCT04321278 |
| Goldman 2020 | Goldman JD, Lye DC, Hui DS, et al. Remdesivir for 5 or 10 days in patients with severe Covid-19. New England Journal of Medicine. 2020 Nov 5;383(19):1827-37. | 10.1056/NEJMoa2015301 | NCT04292899 |
| Guvenmez 2020 | Guvenmez O, Keskin H, Ay B, et al. The comparison of the effectiveness of Lincocin® and Azitro® in the treatment of covid-19-associated pneumonia: A prospective study. Journal of Population Therapeutics and Clinical Pharmacology. 2020 Jun 9;27(SP1):e5-10. | 10.15586/jptcp.v27iSP1.684 | NA |
| Horby 2020a | Horby P, Lim WS, Emberson JR, et al., RECOVERY Collaborative Group. Dexamethasone in hospitalized patients with Covid-19 - preliminary report. N Engl J Med 2021. Feb 25;384(8):693-704 | 10.1056/NEJMoa2021436 | ISRCTN50189673; NCT04381936 |
| Horby 2020b | Horby P, Mafham M, Linsell L, et al. Effect of Hydroxychloroquine in Hospitalized Patients with COVID-19: Preliminary results from a multi-centre, randomized, controlled trial. medRxiv. 2020.07.15.20151852 | 10.1101/2020.07.15.20151852 | ISRCTN50189673; NCT04381936 |
| Horby 2020c | Horby PW, Mafham M, Bell JL, et al. Lopinavir-ritonavir in patients admitted to hospital with COVID-19 (RECOVERY): a randomised, controlled, open-label, platform trial. The Lancet. 2020 Oct 24;396(10259):1345-52. | 10.1016/S0140-6736(20)32013-4 | ISRCTN50189673; NCT04381936 |
| Hu 2020 | Hu K, Wang M, Zhao Y, et al. A small-scale medication of leflunomide as a treatment of COVID-19 in an open-label blank-controlled clinical trial. Virologica Sinica. 2020 Jul 21:1-9. | 10.1007%2Fs12250-020-00258-7 | ChiCTR2000030058 |
| Huang 2020a | Huang M, Tang T, Pang P, et al. Treating COVID-19 with chloroquine. Journal of molecular cell biology. 2020 Apr;12(4):322-5. | 10.1093/jmcb/mjaa014 | ChiCTR2000029542 |
| Huang 2020b | Huang YQ, Tang SQ, Xu XL, et al. No statistically apparent difference in antiviral effectiveness observed among ribavirin plus interferon-alpha, lopinavir/ritonavir plus interferon-alpha, and ribavirin plus lopinavir/ritonavir plus interferon-alpha in patients with mild to moderate coronavirus disease 2019: results of a randomized, open-labeled prospective study. Frontiers in Pharmacology. 2020 Jul 14;11:1071. | 10.3389/fphar.2020.01071 | ChiCTR2000029387 |
| Hung 2020 | Hung IF, Lung KC, Tso EY, et al. Triple combination of interferon beta-1b, lopinavir-ritonavir, and ribavirin in the treatment of patients admitted to hospital with COVID-19: an open-label, randomised, phase 2 trial. The Lancet. 2020 May 30;395(10238):1695-704. | 10.1016/S0140-6736(20)31042-4 | NCT04276688 |
| Ivaschenko 2020 | Ivashchenko AA, Dmitriev KA, Vostokova NV, et al. AVIFAVIR for treatment of patients with moderate COVID-19: interim results of a phase II/III multicenter randomized clinical trial. medRxiv 2020.07.26.20154724 | 10.1101/2020.07.26.20154724 | NCT04434248 |
| Jeronimo 2020 | Jeronimo CM, Farias ME, Val FF, et al. Methylprednisolone as adjunctive therapy for patients hospitalized with COVID-19 (Metcovid): a randomised, double-blind, phase IIb, placebo-controlled trial. Clin Infect Dis. 2021 May 4;72(9):e373-e381 | 10.1093/cid/ciaa1177 | NCT04343729 |
| Kimura 2020 | Kimura KS, Freeman MH, Wessinger BC, et al. Interim analysis of an open-label randomized controlled trial evaluating nasal irrigations in non-hospitalized patients with coronavirus disease 2019. Int Forum Allergy Rhinol. 2020 Dec;10(12):1325-132 | 10.1002/alr.22703 | NA |
| Lemos 2020 | Lemos AC, do Espí­rito Santo DA, Salvetti MC, et al. Therapeutic versus prophylactic anticoagulation for severe COVID-19: A randomized phase II clinical trial (HESACOVID). Thrombosis research. 2020 Dec 1;196:359-66. | 10.1016/j.thromres.2020.09.026 | REBEC RBR-949z6v |
| Li 2020a | Li Y, Xie Z, Lin W, et al. An exploratory randomized, controlled study on the efficacy and safety of lopinavir/ritonavir or arbidol treating adult patients hospitalized with mild/moderate COVID-19 (ELACOI). medRxiv 2020.03.19.20038984 | 10.1101/2020.03.19.20038984 | NCT04252885 |
| Li 2020b | Li C, Luo F, Liu C, Xiong N, et al. Engineered interferon alpha effectively improves clinical outcomes of COVID-19 patients. Research Square https://doi.org/10.21203/rs.3.rs-65224/v1 | 10.21203/rs.3.rs-65224/v1 | ChiCTR2000029638 |
| Li 2020c | Li T, Sun L, Zhang W, Zheng C, et al. Bromhexine hydrochloride tablets for the treatment of moderate COVID-19: an open label randomized controlled pilot study. Clinical and translational science. 2020 Nov;13(6):1096-102. | 10.1111/cts.12881 | NCT04273763 |
| Lopes 2020 | Lopes MI, Bonjorno LP, Giannini MC, et al. Beneficial effects of colchicine for moderate to severe COVID-19: an interim analysis of a randomized, double-blinded, placebo controlled clinical trial. medRxiv. 2020.08.06.20169573 | 10.1101/2020.08.06.20169573 | RBR-8jyhxh |
| Lou 2020 | Lou Y, Liu L, Yao H, et al. Clinical outcomes and plasma concentrations of baloxavir marboxil and favipiravir in COVID-19 patients: An exploratory randomized, controlled trial. European Journal of Pharmaceutical Sciences. 2021 Feb 1;157:105631. | 10.1016/j.ejps.2020.105631 | ChiCTR2000029544 |
| Lyngbakken 2020 | Lyngbakken MN, Berdal JE, Eskesen A, et al. A pragmatic randomized controlled trial reports lack of efficacy of hydroxychloroquine on coronavirus disease 2019 viral kinetics. Nature communications. 2020 Oct 20;11(1):1-6. | 10.1038/s41467-020-19056-6 | NCT04316377 |
| Mansour 2020 | Mansour E, Palma AC, Ulaf RG, et al. Pharmacological inhibition of the kinin-kallikrein system in severe COVID-19 A proof-of-concept study. medRxiv. 2020.08.11.20167353 | 10.1101/2020.08.11.20167353 | U1111-1250-1843 |
| Mehboob 2020 | Mehboob R, Ahmad F, Qayyum A, et al. Aprepitant as a combinant with Dexamethasone reduces the inflammation via Neurokinin 1 Receptor Antagonism in severe to critical Covid-19 patients and potentiates respiratory recovery: A novel therapeutic approach. medRxiv. 2020.08.01.20166678 | 10.1101/2020.08.01.20166678 | NCT04468646 |
| Miller 2020 | Miller J, Bruen C, Schnaus M, et al. Auxora versus standard of care for the treatment of severe or critical COVID-19 pneumonia: results from a randomized controlled trial. Critical Care. 2020 Dec;24(1):1-9. | 10.1186/s13054-020-03220-x | NCT04345614 |
| Mitja 2020a | Mitjà O, Corbacho-Monné M, Ubals M, et al. Hydroxychloroquine for early treatment of adults with mild Covid-19: a randomized-controlled trial. Clinical Infectious Diseases. 2020 Jul 16;ciaa1009. | 10.1093/cid/ciaa1009 | NCT04304053 |
| Mitja 2020b | MitjÃ O, Corbacho M, G-Beiras C, et al. Hydroxychloroquine alone or in combination with cobicistat-boosted darunavir for treatment of mild COVID-19: a cluster-randomized clinical trial. SSRN, 2020. https://ssrn.com/abstract=3615997 | 10.2139/ssrn.3615997 | NCT04304053; EUCTR2020-001031-27 |
| Nojomi 2020 | Nojomi, M., Yassin, Z., Keyvani, H. et al. Effect of Arbidol (Umifenovir) on COVID-19: a randomized controlled trial. BMC Infect Dis. 2020 Dec 14;20(1):954 | 10.1186/s12879-020-05698-w | IRCT20180725040596N2 |
| Pan 2020 | Pan H, Peto R, Abdool Karim Q, Alejandria M, Henao Restrepo AM, Hernandez Garcia C. Repurposed antiviral drugs for COVID-19; interim WHO SOLIDARITY trial results. medRxiv. 2020.10. 15.20209817. | https://doi.org/10.1101/2020.10.15.20209817 | ISRCTN83971151; NCT04315948 |
| Rahmani 2020 | Rahmani H, Davoudi-Monfared E, Nourian A, et al. Interferon ß-1b in treatment of severe COVID-19: A randomized clinical trial. International immunopharmacology. 2020 Nov 1;88:106903. | 10.1016/j.intimp.2020.106903 | IRCT20100228003449N27 |
| Ren 2020 | Ren Z, Luo H, Yu Z, et al. A Randomized, Open Label, Controlled Clinical Trial of Azvudine Tablets in the Treatment of Mild and Common COVID-19, a Pilot Study. Advanced Science. 2020 Oct;7(19):2001435. | 10.1002/advs.202001435 | ChiCTR2000029853 |
| Rosas 2020 | Rosas I, Bräu N, Waters M, et al. Tocilizumab in hospitalized patients with COVID-19 pneumonia. medRxiv. 2020.08.27.20183442 | 10.1101/2020.08.27.20183442 | NCT04320615 |
| Sadeghi 2020 | Sadeghi A, Ali Asgari A, Norouzi A, et al. Sofosbuvir and daclatasvir compared with standard of care in the treatment of patients admitted to hospital with moderate or severe coronavirus infection (COVID-19): a randomized controlled trial. Journal of Antimicrobial Chemotherapy. 2020 Nov;75(11):3379-85. | 10.1093/jac/dkaa334 | IRCT20200128046294N2 |
| Salehzadeh 2020 | Salehzadeh F, Pourfarzi F, Ataei S. The Impact of Colchicine on The COVID-19 Patients; A Clinical Trial Study. Research Square. 2020 https://doi.org/10.21203/rs.3.rs-69374/v1 | 10.21203/rs.3.rs-69374/v1 | IRCT20200418047126N1 |
| Sekhavati 2020 | Sekhavati E, Jafari F, SeyedAlinaghi S, et al. Safety and effectiveness of azithromycin in patients with COVID-19: An open-label randomised trial. International journal of antimicrobial agents. 2020 Oct 1;56(4):106143. | 10.1016/j.ijantimicag.2020.106143 | NA |
| Silva Borba 2020 | Borba MG, Val FF, Sampaio VS, et al. Effect of high vs low doses of chloroquine diphosphate as adjunctive therapy for patients hospitalized with severe acute respiratory syndrome coronavirus 2 (SARS-CoV-2) infection: a randomized clinical trial. JAMA network open. 2020 Apr 1;3(4):e208857-. | 10.1001/jamanetworkopen.2020.8857 | NCT04323527 |
| Skipper 2020 | Skipper CP, Pastick KA, Engen NW, et al. Hydroxychloroquine in nonhospitalized adults with early COVID-19: a randomized trial. Annals of internal medicine. 2020 Oct 20;173(8):623-31. | 10.7326/M20-4207 | NCT04308668 |
| Spinner 2020 | Spinner CD, Gottlieb RL, Criner GJ, et al. Effect of remdesivir vs standard care on clinical status at 11 days in patients with moderate COVID-19: a randomized clinical trial. Jama. 2020 Sep 15;324(11):1048-57. | 10.1001/jama.2020.16349 | NCT04292730 |
| Tang 2020 | Tang W, Cao Z, Han M, et al. Hydroxychloroquine in patients with mainly mild to moderate coronavirus disease 2019: open label, randomised controlled trial.  BMJ. 2020 May 14;369:m1849. | 10.1136/bmj.m1849 | ChiCTR2000029868 |
| Tomazini 2020 | Tomazini BM, Maia IS, Cavalcanti AB, et al. Effect of dexamethasone on days alive and ventilator-free in patients with moderate or severe acute respiratory distress syndrome and COVID-19: the CoDEX randomized clinical trial. Jama. 2020 Oct 6;324(13):1307-16. | 10.1001/jama.2020.17021 | NCT04327401 |
| Ulrich 2020 | Ulrich RJ, Troxel AB, Carmody E, et al. Treating COVID-19 With Hydroxychloroquine (TEACH): A Multicenter, Double-Blind Randomized Controlled Trial in Hospitalized Patients. Open Forum Infect Dis. 2020 Sep 23;7(10):ofaa446 | 10.1093/ofid/ofaa446 | NCT04369742 |
| Vlaar 2020 | Vlaar AP, de Bruin S, Busch M, et al. Anti-C5a antibody IFX-1 (vilobelimab) treatment versus best supportive care for patients with severe COVID-19 (PANAMO): an exploratory, open-label, phase 2 randomised controlled trial. The Lancet Rheumatology. 2020 Dec 1;2(12):e764-73. | 10.1016/S2665-9913(20)30341-6 | NCT04333420 |
| Wang 2020a | Wang Y, Zhang D, Du G, et al. Remdesivir in adults with severe COVID-19: a randomised, double-blind, placebo-controlled, multicentre trial. The Lancet. 2020 May 16;395(10236):1569-78. | 10.1016/S0140-6736(20)31022-9 | NCT04257656 |
| Wang 2020c | Wang SZ, Wang HJ, Chen HM, et al. Lianhua Qingwen capsule and interferon-Î± combined with lopinavir/ritonavir for the treatment of 30 COVID-19 patients. J Bengbu Med Coll. 2020;45(2):154-5. | 10.13898/j.cnki.issn.1000-2200.2020.02.004 | NA |
| Wang 2020d | Wang D, Fu B, Peng Z, et al. Tocilizumab ameliorates the hypoxia in COVID-19 moderate patients with bilateral pulmonary lesions: a randomized, controlled, open-label, multicenter trial. | 10.2139/ssrn.3667681 | ChiCTR2000029765 |
| Wang 2020e | Wang M, Zhao Y, Hu W, et al. Treatment of COVID-19 Patients with Prolonged Post-Symptomatic Viral Shedding with Leflunomide--a Single-Center, Randomized, Controlled Clinical Trial. Clin Infect Dis. 2020 Sep 21;ciaa1417 | 10.1093/cid/ciaa1417 | ChiCTR2000030058 |
| Wu 2020 | Wu X, Yu K, Wang Y, et al. Efficacy and safety of triazavirin therapy for coronavirus disease 2019: A pilot randomized controlled trial. Engineering. 2020 Oct 1;6(10):1185-91. | 10.1016/j.eng.2020.08.011 | ChiCTR20000300001 |
| Yethindra 2020 | Yethindra V, Tagaev T, Uulu MS, Parihar Y. Efficacy of umifenovir in the treatment of mild and moderate COVID-19 patients. International Journal of Research in Pharmaceutical Sciences. 2020 Aug 19;11:506-9. | 10.26452/ijrps.v11ispl1.2839 | NA |
| Yuan 2020 | Yuan X, Yi W, Liu B, et al. Pulmonary radiological change of COVID-19 patients with 99mTc-MDP treatment. medRxiv. 2020.04.07.20054767 | 10.1101/2020.04.07.20054767 | CHICTR2000029431 |
| Zhang 2020 | Zhang J, Rao X, Li Y, et al. High-dose vitamin C infusion for the treatment of critically ill COVID-19. Research Square. 2020. https://doi.org/10.21203/rs.3.rs-52778/v2 | 10.21203/rs.3.rs-52778/v2 | NCT04264533 |
| Zhao 2020 | Zhao H, Zhu Q, Zhang C, et al. Tocilizumab combined with favipiravir in the treatment of COVID-19: A multicenter trial in a small sample size. Biomedicine & Pharmacotherapy. 2021 Jan 1;133:110825. | 10.1016/j.biopha.2020.110825 | ChiCTR2000030894; NCT04310228 |
| Zheng 2020 | Zheng F, Zhou Y, Zhou Z, et al. SARS-CoV-2 clearance in COVID-19 patients with Novaferon treatment: A randomized, open-label, parallel-group trial. International Journal of Infectious Diseases. 2020 Oct 1;99:84-91. | 10.1016/j.ijid.2020.07.053 | ChiCTR2000029496 |
| Zhong 2020 | Zhong M, Sun A, Xiao T, et al. A randomized, single-blind, group sequential, active-controlled study to evaluate the clinical efficacy and safety of Î±-lipoic acid for critically ill patients with coronavirus disease 2019 (COVID-19). medRxiv 2020.04.15.20066266 | 10.1101/2020.04.15.20066266 | ChiCTR2000029851 |
| de Alencar 2020 | de Alencar JC, Moreira CD, Müller AD, et al. Double-blind, Randomized, Placebo-controlled Trial With N-acetylcysteine for Treatment of Severe Acute Respiratory Syndrome Caused by Coronavirus Disease 2019 (COVID-19). Clin Infect Dis. 2020 Sep 23;ciaa1443 | 10.1093/cid/ciaa1443 | RBR-8969zg; U1111-1250-356 |
