## Supplementary tables and figures for "Definitions matter: heterogeneity of COVID-19 disease severity criteria and incomplete reporting compromise meta-analysis"

**Supplementary Table 3.** Agreement between WHO - COVID-19 Living Network Meta-Analysis and COVID-NMA initiative groups with respect to possible minimum and maximum severity of trial participants (n=70)

|  | **Discordant maximum severity** | **Concordant maximum severity** |
| --- | --- | --- |
| **Discordant minimum severity** | 14 (20%) | 15 (21%) |
| **Concordant minimum severity** | 15 (21%) | 26 (37%) |
