## Supplementary tables and figures for "Definitions matter: heterogeneity of COVID-19 disease severity criteria and incomplete reporting compromise meta-analysis"

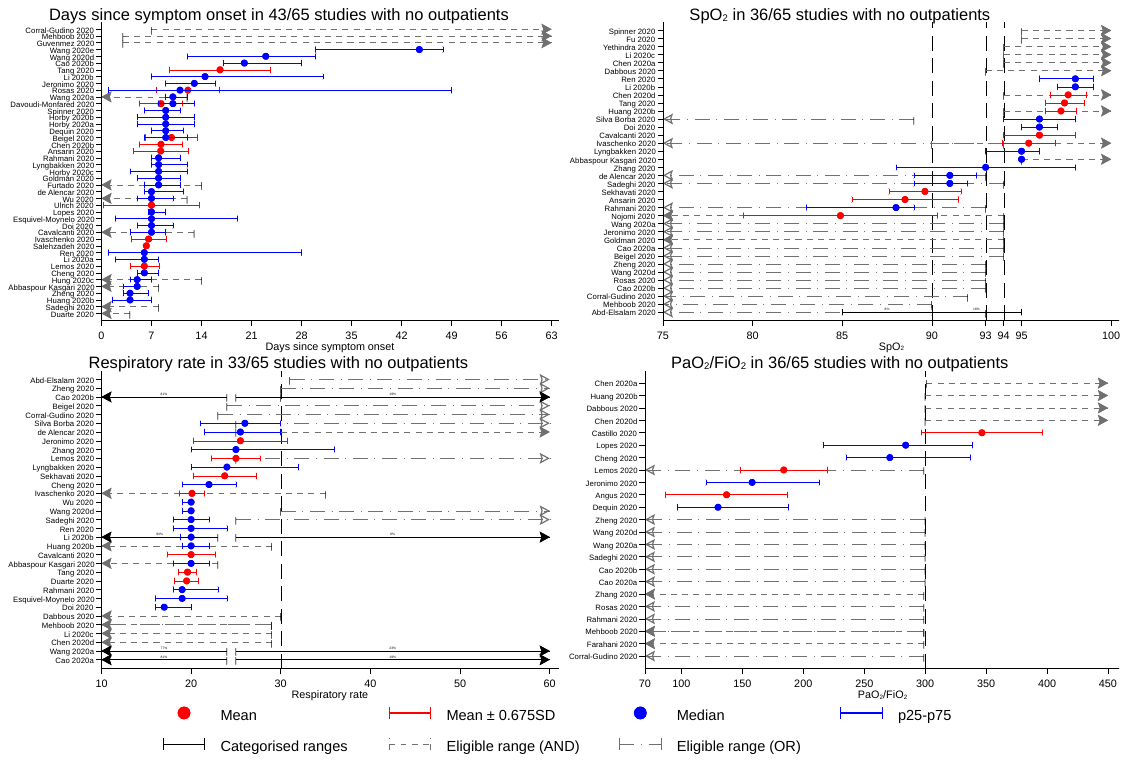

Supplementary Figure 2 No outpatients subset - Available information on days since symptom onset; SpO_2_; respiratory rate; PaO_2_/FiO_2_. Dashed vertical lines denote thresholds used by either the NIH; WHO; Covid-NMA or Magic NMA groups to categorise severity. Eligible range (AND) refers to a criterion that is part of an AND condition and therefore an individual falling into this range would also require at least one other criterion to be met; eligible range (OR) refers to a criterion that is part of an OR condition.
